# Visual outcome and associated risk factors in patients following small incision cataract surgery at the National Referral Hospital, Bhutan

**DOI:** 10.1101/2024.11.23.24317844

**Authors:** Sandip Tamang, Nor Tshering Lepcha, Mendu Dukpa, Karma Tenzin

## Abstract

**Purpose:** To evaluate small incision cataract surgery visual outcomes and associated risk factors in a tertiary eye care center in Bhutan.

**Methods:** This was a hospital-based prospective longitudinal study. A total of 310 patients who met the eligibility criteria were included in the study.

**Results:** The mean age of the patients was 68.36 (SD: 12.74 years), and 157 (50.6%) were male. 285 (91.94%) of the cataract-operated eyes had best-corrected vision greater than 6/60, while 25 (8.06%) of the eyes had vision less than 6/60. 47 individuals (15.16%) had pre-existing ocular co-morbidity. Intraoperative complications were seen in 19 eyes (6%). At the 6-week follow-up visit, based on best-corrected visual acuity (BCVA), 90.65% had a good visual outcome. Analysis of multiple logistic regression showed that patients with preoperative ocular comorbidities (OR 50.92; 95% CI 10.23, 253.37) and those with operative complications (OR 16.59; 95% CI 3.54, 77.70) were significantly associated with poor visual outcome.

**Conclusions:** This study shows that cataract surgery can restore good visual acuity. However, patients should be followed up on for a longer period, and greater emphasis should be given to monitoring outcomes after cataract surgery.

## Introduction

The World Health Organization (WHO) reports that globally, at least 1 billion people have preventable near or distance vision impairment. Cataract is the leading cause of vision impairment and blindness in 65.2 million people globally (1). In Southeast Asia, the proportion of blindness due to cataracts was 44% (2).

Bhutan, located in the Eastern Himalayas between India and China, has a population of 7, 35,553 (3). The prevalence of visual impairment in Bhutan was 1%, with cataracts contributing as the major cause (48.3%) of preventable blindness.

A cataract is a clouding of the natural intraocular lens and, if not corrected can lead to diminution of vision and irreversible blindness. Small incision cataract surgery (SICS) involves the removal of a cloudy lens and the implantation of an intraocular lens (IOL) (4). It is the most effective method to restore vision.

However, the outcome of the cataract surgery determines the vision-related functioning and improvement in the patient’s life, which is measured in terms of visual acuity. WHO recommends more than 80% of eyes with available correction between discharge and 12 weeks postoperatively should have good visual acuity of 6/6-6/18, and less than 5% of eyes should have poor visual acuity of <6/60 (5).

However, the Rapid Assessment of Avoidable Blindness (RAAB) survey conducted in 2018 noted a postoperative good visual outcome of 67.3% for the country, which was found to be below the WHO recommendation (6).

Studies have shown that increasing patient age is associated with poor visual outcomes following SICS. Studies conducted in Bangladesh showed that the risk for poor visual outcomes was increased by 4.63-fold among patients aged 70 years and above (7). Studies in Liberia and India also reported similar findings with older age (8–10). One of the large studies conducted in 4 sites in North America and Europe, which included the United States, Spain, Canada, and Denmark, also demonstrated that older age was associated with poor visual outcomes (11).

The second RAAB survey conducted in the country in 2018 found that following SICS, the main cause of poor visual outcome was ocular comorbidities in 43.6%. These included corneal opacities, glaucoma, and posterior segment diseases such as diabetic retinopathy and age-related macular degeneration (6). The presence of pre-existing ocular pathology is a strong independent predictor of visual outcomes after SICS, as found in a study in India (12). Other studies in India have also shown that poor visual outcomes were due to pre-existing ocular comorbidities impacting the visual functioning of the patients (13, 14). Visual outcomes of SICS in Denmark, the United States, Spain, and Canada showed poorer outcomes with coexisting ocular comorbidities (11). Similar findings have also been noted in studies conducted in Australia and Nigeria where ocular comorbidities affected the final visual outcome of the patients (15–17). A study in Nepal by Hening et al. also demonstrated poor visual outcomes in those patients who had ocular comorbidities (18).

The RAAB survey conducted for the country in 2018 noted that surgical complications amounted to 40.5 %, while postoperative sequelae amounted to 22.2%, which impacted the final visual outcomes of the patients resulting in the good visual outcomes being below WHO recommendation for the country (6).

Cox JT et al. noted the mean SICS complication rate for all surgeons to be 1.7±1.0% for both intraoperative and postoperative complications (9).

A study in Nepal demonstrated that postoperative complications, mainly the postoperative sequelae such as PCO, cause poor visual outcomes in the patients (19). Another study in Nepal also showed that surgical complications such as posterior capsular rent vitreous loss and endophthalmitis that occurred caused poor visual outcomes (20). Ruit S et al. also inferred a relation between poor visual outcomes and surgical compilations (21).

Studies conducted in African countries had good postoperative visual acuity below the WHO recommended criteria, indicating a similar pattern for developing countries (22, 23). Norregaard JC et al. demonstrated a superior visual outcome following SICS above the WHO recommendation, which was conducted in the United States, Canada, Denmark, and Spain (11). A similar outcome has also been noted in studies in India (10). However, there are studies in Bangladesh and Nigeria that reported poor outcomes following cataract surgeries (7, 16).

In Bhutan, Jigme Dorji Wangchuck National Referral Hospital (JDWNRH) functions as the only tertiary-level eye care center in the country, and there is a need to routinely monitor the outcome of cataract surgery and to follow standardized cataract surgical protocols and postoperative follow-up care.

The study aimed to evaluate postoperative visual acuity and study factors associated with poor visual outcomes and form the baseline data for the Department of Ophthalmology, JDWNRH.

## Material and Methods

This is a hospital-based study conducted at the Gyalyum Kesang Choden Wangchuck National Eye Center (GKCWNEC), Jigme Dorji Wangchuck National Referral Hospital (JDWNRH), Bhutan. The study design was a prospective longitudinal study. Patients who had visited the GKCWNEC and had undergone small incision cataract surgery (SICS) from 1st September 2021 to 30th April 2022 were eligible to be included in this study. The sampling method was census sampling. Patients who were 18 years and above were included. Consequently, congenital cataracts, patients with secondary intraocular (IOL) implantation, and those undergoing combined procedures with SICS (e.g., SICS with corneal graft or SICS with trabeculectomy, etc.) were excluded. Due to poor visual prognosis after surgery, these patients were excluded.

All socio-demographic data were collected from the patients, and the pre-operative and post-operative data were collected from the patient’s medical record after following up with the patient, which was recorded in the standard cataract surgical record (CSR) form. The standard pre-operative examination included a detailed history, measurements of presenting visual acuity (PVA), and best corrected visual acuity (BCVA) with Snellen charts. Computerized Snellen charts were used to test PVA and BCVA. The visual acuity of each eye was classified according to the WHO-recommended guideline for cataract outcome grading. PVA and BCVA were measured by optometrists in refraction rooms where computerized Snellen charts were used, which measured the presenting visual acuity without correction and best corrected visual acuity with pinhole. All these records were directly obtained from the patient’s documents which were recorded before SICS and then at day 1, day 7, and 6 weeks post-SICS. Intraocular pressure measurement was done with an Icare (rebound tonometer) as well as applanation tonometry where needed.

Patients were assessed by ophthalmologists in different consultation chambers before SICS and then followed up and assessed on day 1, day 7, and 6 weeks post-SICS. The patients had detailed slit lamp assessment including anterior segment examination of cornea, anterior chamber, and pupils and dilated examination to assess the lens status which was classified according to Lens Opacity Classification System III. Posterior segment pathology was assessed with a +90D lens. Any ocular pathology (anterior or posterior segment) was noted if present. Ocular comorbidities were grouped as corneal scars, old iritis, retinal diseases, glaucoma, and others. All the findings were noted in the patient’s documents and were obtained when the patient presented to the pre-operative chamber. Before the surgery, biometry and sac syringing was done for all the patients and they also underwent blood investigations (viral markers, TPHA (Treponema Pallidum Hemagglutination Assay), RPR (Rapid Plasma Reagin) and random blood sugar). Any findings that were either not clear or missing were directly consulted with the assessing ophthalmologist and recorded in the CSR. Standard Operating Procedures (SOP) and definitions specified in the World Health Organization (WHO) Manual for Monitoring Cataract Surgical Outcomes (MCSO) were followed. For assessment of visual outcome, visual acuities were categorized according to WHO guidelines: good (6/6-6/18), borderline (6/24-6/60), or poor surgical outcomes (<6/60).

The study questionnaire was coded and a data documentation sheet was prepared. Data were double-entered between September 2021 and June 2022, validated, and analyzed using EpiData (version 3.1 for entry and version 2.2.2.183 for analysis, EpiData Association, Odense, Denmark). Additional analysis was performed using STATA (version 13.0, licensed by KGUMSB, Bhutan). Factors associated with visual acuity (age, ocular morbidity, systemic comorbidity, postoperative complications) were tested using unadjusted analysis using logistic regression. Those factors with p < 0.1 were included in the final model for adjusted analysis. For all statistical analysis, p-value < 0.05 was considered significant.

The Ethics Committee of the Khesar Gyalpo University of Medical Sciences of Bhutan (KGUMSB), Bhutan, approved this study and it was conducted according to the tenets of the Declaration of Helsinki.

## Results

### Demographic characteristics

From 1st September 2021 to 30th April 2022, 430 patients underwent SICS, out of which 372 were included in the study and fulfilled the inclusion criteria. Among that 310 patients completed the follow-up with a response rate of 83%, while 62 were lost to follow-up. These 62 patients who were lost to follow-up were all living outside Thimphu and did not have their follow-up at the Department of Ophthalmology, JDWNRH. The remaining 310 patients were then analyzed. Amongst the 310 patients analyzed in the study, there were 157 male and 153 female patients. The mean age among the group was 68.36 (SD 12.74).

As illustrated in Figure 1, the highest percentage [70.65%, n = 219] of the patients belonged to the age group above 65 years followed by the age group of 55 – 65 years [16.77%, n= 52]. Amongst the patients analyzed in the study, 74.52% (n=231) had no schooling. Unemployed patients accounted for 94.19% (n=292) while patients living outside Thimphu accounted for 87.42% (n=271). Table 1 provides a breakdown of the qualification, employment status, and present address.

**Figure 1:**
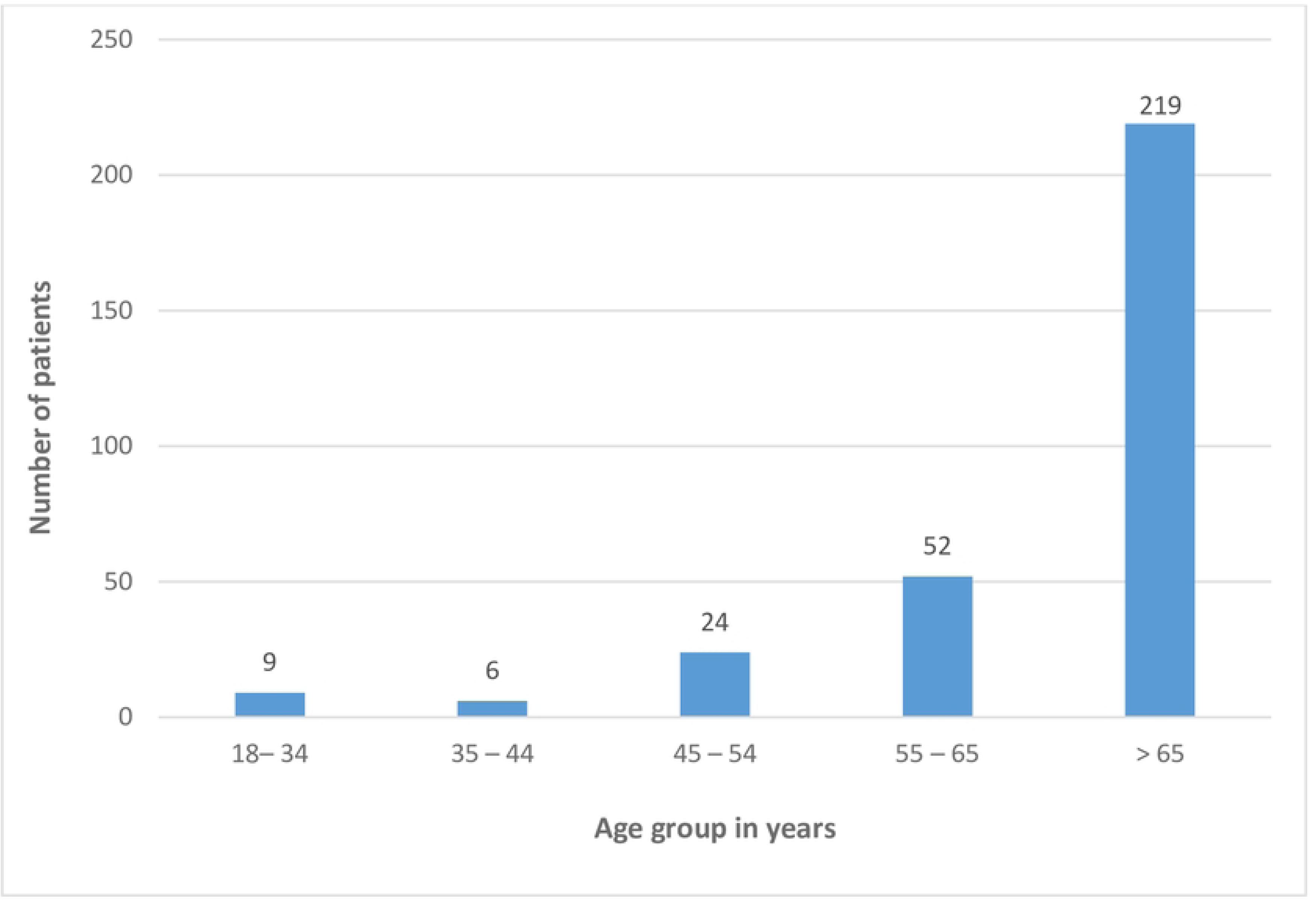
Age distribution of patients who underwent SICS at Jigme Dorji Wangchuck National Referral Hospital, Thimphu, Bhutan from 1^st^ September 2021 to 30^th^ April 2022 (n=310)

**Table 1:**
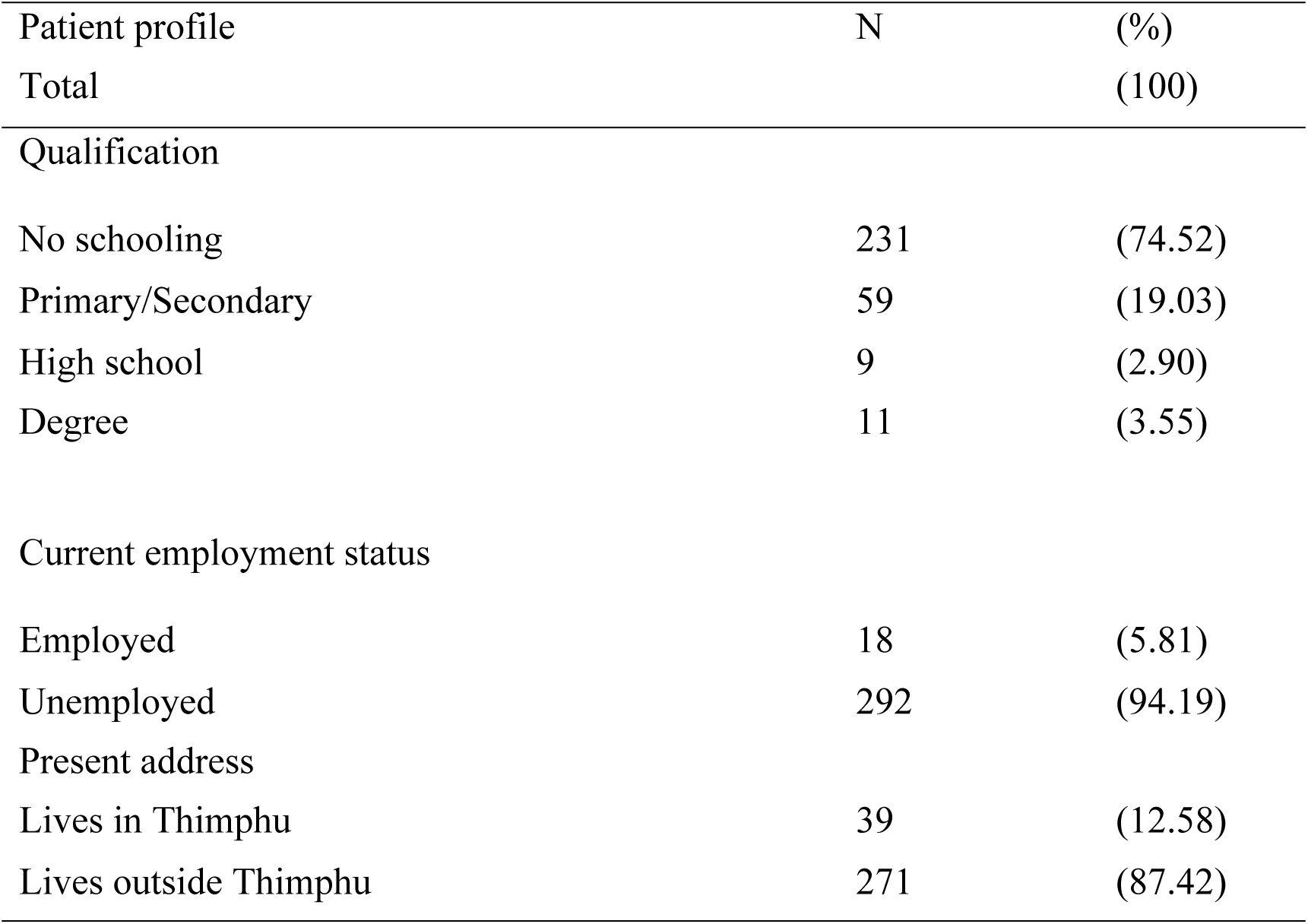
Employment status, qualification, and present address of the patients who underwent SICS at Jigme Dorji Wangchuck National Referral Hospital, Thimphu, Bhutan from 1^st^ September 2021 to 30^th^ April 2022 (n=310)

According to the occupation of the patients, the maximum were farmers (n=173) followed by housewives (n=107) and the rest were in other categories as depicted in Figure 2. Socio-demographic profiles such as age, gender, working status, and occupation associated with poor visual outcomes were analyzed and it was found to have no significant association (p-value >0.05) as depicted in Table 2.

**Figure 2:**
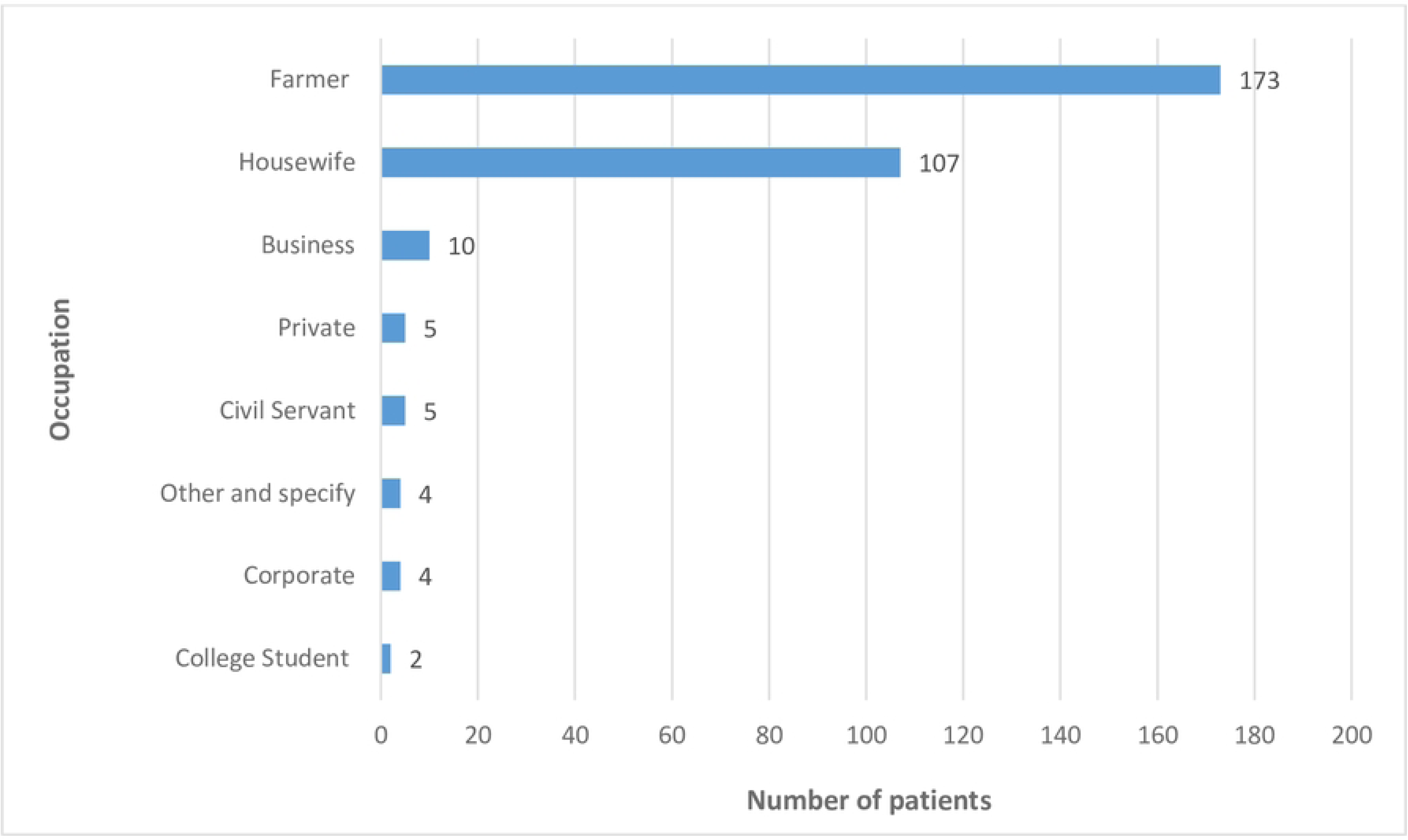
Occupation of patients who underwent SICS at Jigme Dorji Wangchuck National Referral Hospital, Thimphu, Bhutan from 1^st^ September 2021 to 30^th^ April 2022 (n=310)

**Table 2:** Socio-demographic factors associated with poor visual outcome after SICS at Jigme Dorji Wangchuck National Referral Hospital from 1^st^ September 2021 to 30^th^ April 2022.

| Socio-demographic characteristics |  | N | (%) | Unadjusted OR | aOR | 95% CI | p-value |
| --- | --- | --- | --- | --- | --- | --- | --- |
| Age group |  |  |  |  |  |  |  |
|  | 18– 34 | 9 | (2.90) | Reference | Reference |  |  |
|  | 35 – 44 | 6 | (1.94) | 2.50 | 2.95 | 0.20-42.83 | 0.427 |
|  | 45 – 54 | 24 | (7.74) | 11.50 | 11.31 | 0.92-138.54 | 0.058 |
|  | 55 – 65 | 52 | (16.77) | 12.50 | 6.73 | 0.75-60.03 | 0.088 |
|  | Above 65 | 219 | (70.65) | 5.58 | 2.73 | 0.43-17.03 | 0.281 |
| Gender |  |  |  |  |  |  |  |
| Male |  | 157 | (50.65) | 1.33 |  |  |  |
| Female |  | 153 | (49.35) | Reference |  |  |  |
| Working status |  |  |  |  |  |  |  |
| Employed |  | 18 | (5.81) | 0.27 | 4.13 | 0.03-1.40 | 0.113 |
| Not employed |  | 292 | (94.19) | Reference | Reference |  |  |
| Occupation |  |  |  |  |  |  |  |
| Civil Servant/ Private/ Corporate |  | 14 | (4.52) | 0.24 |  |  |  |
| Housewife |  | 107 | (34.52) | 0.82 |  |  |  |
| Farmer |  | 173 | (55.81) | 0.82 |  |  |  |
| Others |  | 16 | (5.16) | Reference |  |  |  |

Among the patients who underwent cataract surgery, nearly 53% (n = 163) had their right eye operated, whereas around 47% (n = 147) had their left eye operated.

Among the patients who underwent cataract surgery, almost 72% (n = 224) had no comorbidities, whereas around 28% had one or more comorbidities (hypertension, diabetes, rheumatoid arthritis, bronchial asthma, syphilis, and chronic obstructive pulmonary disease), as shown in Figure 3.

**Figure 3:**
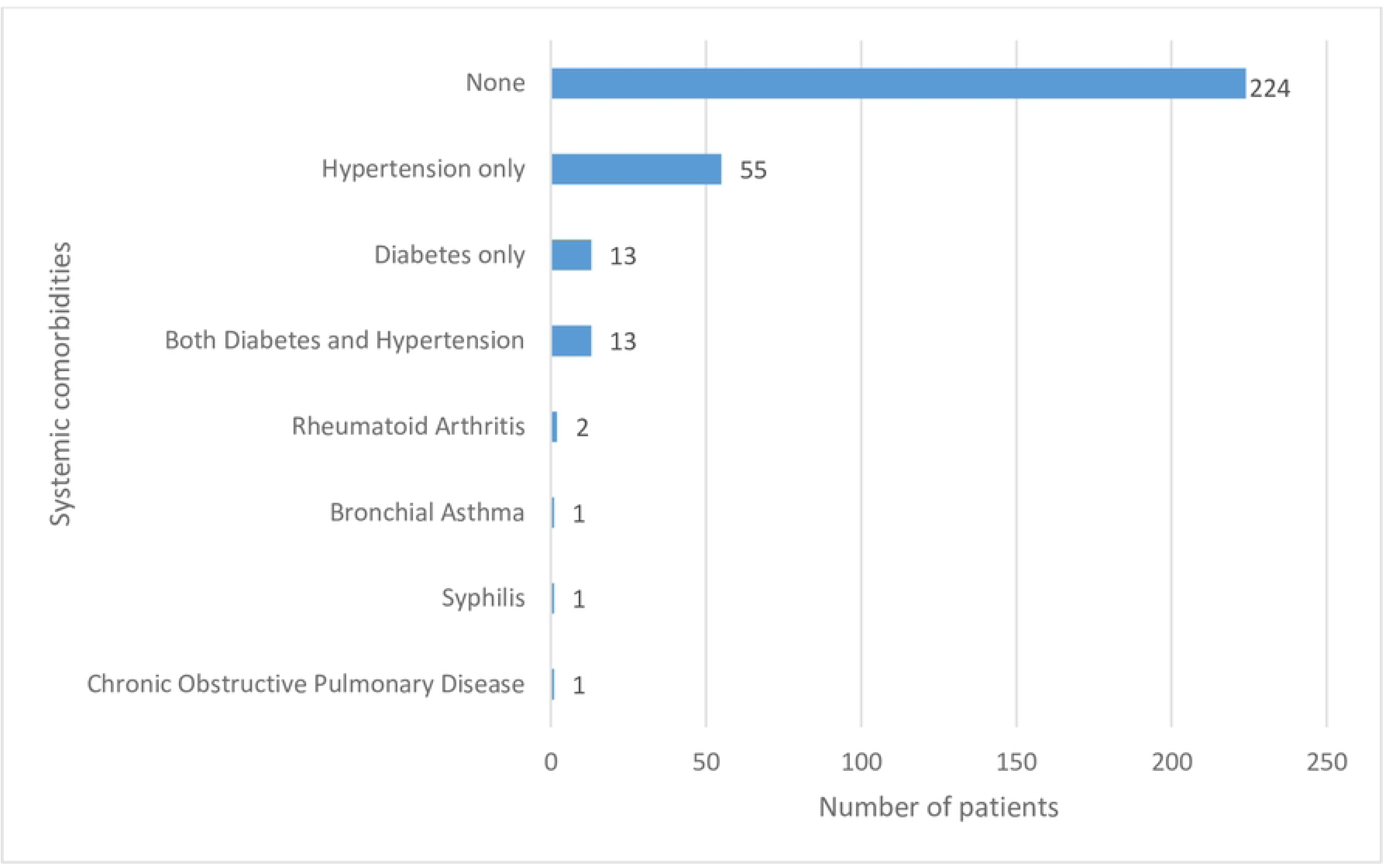
Systemic comorbidities among patients who underwent SICS at Jigme Dorji Wangchuck National Referral Hospital, Thimphu, Bhutan from 1^st^ September 2021 to 30^th^ April 2022 (n=310)

### Visual Acuity Outcome

The visual outcome has been categorized into presenting visual acuity (PVA) and best corrected visual acuity (BCVA) and measured at postoperative day 1, day 7, and 6 weeks. Figure 4 depicts the total number of patients with BCVA at different post-operative visits. Table 3 shows the BCVA before SICS and 6 weeks post-SICS in line with the WHO monitoring of cataract surgical outcomes.

**Figure 4:**
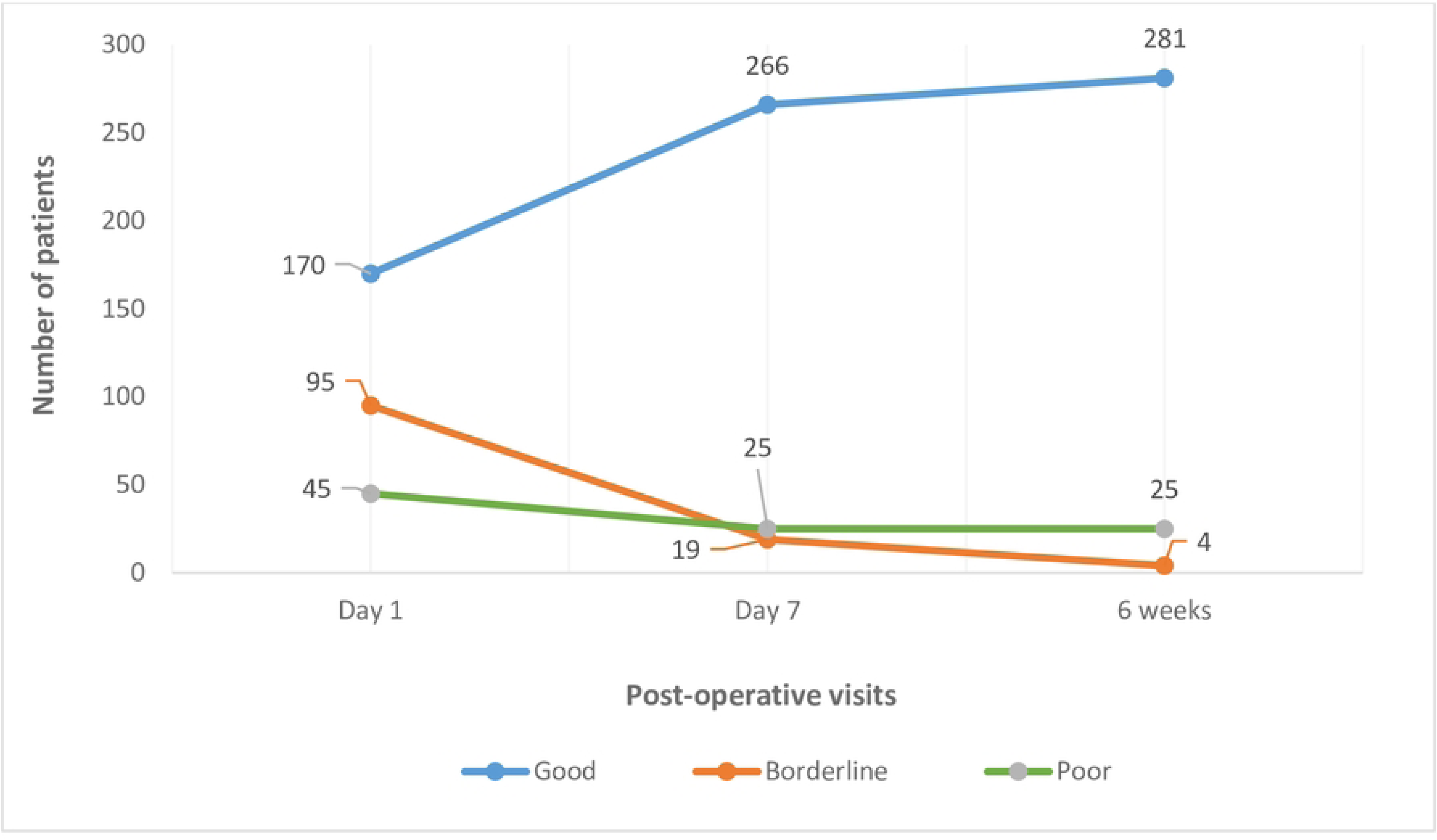
Total number of patients with BCVA at different post-operative visits at Jigme Dorji Wangchuck National Referral Hospital, Thimphu, Bhutan from 1^st^ September 2021 to 30^th^ April 2022 (n=310)

**Table 3:** Best Corrected Visual acuity outcome in the operated eye of the patients before SICS and 6 weeks post-SICS at Jigme Dorji Wangchuck National Referral Hospital, Thimphu, Bhutan from 1^st^ September 2021 to 30^th^ April 2022 (n=310) according to WHO recommendation

| Category of Outcome | Level of VA | Best corrected VA before SICS |  | Best corrected VA at 6 weeks post- SICS |  |
| --- | --- | --- | --- | --- | --- |
|  |  | N | % | N | % |
| Good | 6/6-6/18 | 30 | (9.68) | 281 | (90.65) |
| Borderline | <6/18-6/60 | 125 | (40.32) | 4 | (1.29) |
| Poor | <6/60 | 155 | (50.00) | 25 | (8.06) |

### Ocular pathology, Operative complication, and systemic comorbidities

Ocular pathology was seen in 15.16% (n = 47) of the patients, while 84.84% (n = 263) did not have any ocular pathology. These included corneal scars, retinal disease, glaucoma, and others. From the ocular pathology, a majority had retinal disease (n = 21), followed by others (n = 15), corneal scar (n = 6), and glaucoma (n = 5). Retinal diseases included age-related macular degeneration (ARMD), macular scars, macular holes, and retinal detachment surgery. Other ocular pathologies included complicated uveitic cataracts, retinitis pigmentosa, pseudoexfoliation, traumatic cataracts, Fuchs heterochromic iridocyclitis, optic atrophy, and vitreous hemorrhage. Poor visual outcome was significantly associated with retinal disease (aOR 50.92; 95% CI: 10.23, 253.37), glaucoma (aOR 13.75; 95% CI: 1.26, 150.06), corneal scar (aOR 23.55; 95% CI: 3.01, 184.02), and others (aOR 12.14; 95% CI: 2.36, 62.33), as shown in Table 4.

**Table 4:** Clinical factors associated with poor visual outcome after SICS at Jigme Dorji Wangchuck National Referral Hospital, Thimphu, Bhutan from 1^st^ September 2021 to 30^th^ April 2022.

| Clinical characteristics | N | (%) | Crude OR | aOR | P Value | 95% CI |  |
| --- | --- | --- | --- | --- | --- | --- | --- |
|  |  |  |  |  |  | Lower | Upper |
| Ocular pathology |  |  |  |  |  |  |  |
| Corneal scar | 6 | (1.94) | 25.3 | 23.55 | 0.003 | 3.01 | 184.02 |
| Retinal disease | 21 | (6.77) | 23 | 50.92 | <0.001 | 10.23 | 253.37 |
| Glaucoma | 5 | (1.61) | 6.32 | 13.75 | 0.032 | 1.26 | 150.06 |
| Others | 15 | (4.84) | 12.65 | 12.14 | 0.003 | 2.36 | 62.33 |
| None | 263 | (84.84) | Reference | Reference |  |  |  |
| Operative complications |  |  |  |  |  |  |  |
| None | 291 | (93.87) | Reference | Reference |  |  |  |
| Striate keratopathy | 13 | (4.19) | 9.47 | 16.59 | <0.001 | 3.54 | 77.70 |
| *Others | 6 | (21.94) | - |  |  |  |  |
| Systemic comorbidities |  |  |  |  |  |  |  |
| None | 224 | (72.26) | Reference | Reference |  |  |  |
| Diabetes | 13 | (4.19) | 8.12 | 0.78 | 0.794 | 0.122 | 4.96 |
| Hypertension | 55 | (17.74) | 1.30 | 1.16 | 0.822 | 0.30 | 4.46 |
| Both Diabetes and Hypertension | 13 | (4.19) | 2.36 | 0.81 | 0.847 | 0.09 | 6.76 |
| Others | 5 | (1.61) | 3.25 | - |  |  |  |

Almost 94% (n = 291) had no operative complications, while 6% (n = 19) had operative complications, which included striate keratopathy (n = 13) and others (n = 6) (capsule rupture without vitreous loss, capsule rupture with vitreous loss, and wound leak). Complications like zonular dehiscence, retained lens matter, and endophthalmitis did not occur. Striate keratopathy was mostly seen in patients (n = 13) who already had ocular pathology, and it persisted over time and impacted the visual acuity. It was found to have a significant association with poor visual outcomes (aOR 16.59; 95% CI: 3.54, 77.70).

Systemic comorbidities comprised diabetes, hypertension, and others (such as bronchial asthma, syphilis, chronic obstructive pulmonary disease (COPD), and rheumatoid arthritis). Around 72.26% (n = 224) had no comorbidities, while 27.74% (n = 86) had either one or more systemic illnesses, which did not have any significant association with poor visual outcomes. As very few patients who underwent SICS had vision < 6/60 (n = 25), the causes were mostly selection, amounting to 64% (n = 16), followed by surgery with almost 28% (n = 7) and 8% (n=2) for sequelae (which included posterior capsular opacification (PCO)).

## Discussion

### Demographic characteristics

#### Age

The age distribution was found to be such that the majority were above the age of 65 years, with a mean age of 68.36 ± 12.74 years. The high percentage of patients belonging to the age group above 65 years is consistent with the well-established fact that cataracts primarily affect older individuals (24). In this study, there was no association found between increasing age and the visual outcomes, which is quite contrary to some of the studies that demonstrated a correlation between the two (25, 26)

#### Gender

In this study, there was no predilection to any gender and it was found to be almost similar with 50.65% male and 49.35% female patients.

This study did not find any significant association between gender and final visual outcomes, which is also consistent with several studies in similar contexts (27, 28). It differed from studies where there was a significant association between male gender and improved visual outcome.

This association aligns with several studies (10, 13, 22, 26). Interestingly, some studies in India have shown that the female gender significantly improved postoperative visual acuity, which might be due to improved postoperative self-care among women (29, 30). The relationship between gender and visual acuity is complex and multifaceted considering various contradictory findings.

### Employment status, qualification, and present address

This study found that most of the patients (74.52%) had no schooling and had undergone cataract surgery. It was comparable to a study in Bangladesh where most of the patients were illiterate and most of them lived in rural areas, similar to our study (7). The fact that a significant portion of patients lived outside Thimphu suggests that people from different regions have access to specialized eye care services in the capital city. It can also be noted that the patients who were lost to follow-up were living outside Thimphu and they were not able to return for a postoperative visit at the Department of Ophthalmology, JDWNRH. Since eye care services are provided throughout the country at referral hospitals as well as at the district hospitals, it could be inferred that those lost to follow-up patients were likely seen at one of the health centers.

### Systemic comorbidities

In this study around 72% of the patients had no comorbidities while 28% had one or more comorbidities which consisted of Diabetes, Hypertension, Rheumatoid arthritis, Bronchial asthma, Syphilis, and COPD. The study revealed that the majority of patients had no systemic comorbidities, with diabetes and hypertension being the most prevalent among those who did. These findings are consistent with the high prevalence of diabetes and hypertension in many populations worldwide (31). The absence of a significant association between systemic comorbidities and visual outcomes aligns with some studies (32, 33).

### Ocular comorbidities

The study found a significant association between ocular comorbidities and visual outcomes. Retinal disease amounted to about 44.6% (n=21) of the total ocular comorbidities. Retinal diseases included age-related macular degeneration (ARMD), macular scar, macular hole, and retinal detachment surgery. The patients who had retinal diseases before the surgery had significantly poor visual outcomes. Poor visual outcome was significantly associated with retinal disease (aOR 50.92; 95% CI: 10.23, 253.37), glaucoma (aOR 13.75; 95% CI: 1.26, 150.06), corneal scar (aOR 23.55; 95% CI: 3.01, 184.02), and others (aOR 12.14; 95% CI: 2.36, 62.33). Other ocular pathologies included complicated uveitic cataracts, retinitis pigmentosa, pseudoexfoliation, traumatic cataracts, Fuchs heterochromic iridocyclitis, optic atrophy, and vitreous hemorrhage. The presence of ocular pathology in 15% of the patients is in line with several studies reporting varying rates of ocular comorbidities among cataract patients (34, 35). It was noted that the ocular pathology contributed to poor visual outcomes despite the surgeons performing good SICS. Studies have also noted similar findings where the final visual outcome was affected by the ocular pathology (10, 16, 23, 32). Ocular pathology was found to have a significant effect on the overall visual acuity where it was important to recognize the condition before surgery.

It was similarly noted in the RAAB survey where 43.6% were due to ocular co-morbidities that caused poor visual outcomes. It was also noted that posterior segment diseases such as glaucoma, diabetic retinopathy, and age-related macular degeneration contributed to 15.4% of all visual impairment and 26.1% of blindness (6).

This shows that to detect ocular comorbidities that are likely to affect results, a thorough preoperative eye examination by an ophthalmologist would be required. Then, patients who have ocular comorbidities can be counseled and given a thorough explanation of the outcome of the surgery.

### Postoperative complications

The occurrence of operative complications in 6% of the patients, primarily striate keratopathy, was within the range reported in other studies investigating complications following cataract surgery (13, 36). The study found a significant association between striate keratopathy and visual outcome (aOR 16.59: 95% CI 3.54, 77.70). It was noted that striate keratopathy occurred in all the patients with corneal scar (n=6) demonstrating that the patients with ocular comorbidity such as corneal scar were already at the risk of developing operative complications. So, these patients had poor outcomes which was related to the preexisting ocular pathology. Similar associations were not noted in other studies.

Postoperative complications that were noted in this study were also demonstrated in several other studies that had affected the final visual outcome (14, 30, 37).

However, the absence of severe complications such as zonular dehiscence, retained lens matter, and endophthalmitis was reassuring and suggested a satisfactory safety profile of SICS in our study population. It would be due to experienced surgeons performing the cataract surgeries instead of the resident doctors. It could also be attributed to the experience of the surgeons managing intra-operative complications. Better operative facilities with proper surgical instruments and well-trained operating assistants were other factors that resulted in fewer complications.

### Visual outcomes

According to the WHO and the International Agency for the Prevention of Blindness (IAPB) action plan, good postoperative visual acuity should be obtained in >80% with available correction and >90% with the best correction for a vision of 6/6 - 6/18 (5). The preoperative best corrected good and poor visual outcome was 9.68% (n=30) and 50% (n=155) respectively. The study found that a good visual outcome after 6 weeks of SICS was 90.65% (n=281). It was found to be above the WHO recommendation for monitoring cataract outcomes. This was consistent with the expected positive outcomes of cataract surgery in improving vision (38). It was comparable to studies which had conducted in India and other countries (10, 13, 39–41). This was better than the studies conducted in some developing countries where the visual outcomes were below the WHO recommendation (42–45). The improvement was mainly due to well-trained surgeons and operating assistants, better biometry services, and well-trained staff conducting biometry before the surgery.

However, poor visual outcome was 8.06% (n=25) 6 weeks post-SICS which was more than the WHO recommendation (<5%). This might have resulted due to ocular morbidities as well as surgical complications that had affected the final visual outcome.

It was found that only around 72.7% had good visual outcomes with the best correction in the second RAAB survey, while poor visual outcome was reduced from 15% to 11.5%.

The improvement in the good visual outcome compared to 2018 in its RAAB survey, would be likely due to better healthcare infrastructure in the capital with the shifting of the department into a separate well-established National Eye Care Center with improved facilities and availability of specialized services including vitreoretinal services (60).

### Causes of presenting vision <6/60

The causes of vision <6/60 at the postoperative 6 weeks were found to be majority due to selection (64%, n=16) followed by surgery (28%, n=7) and sequelae (8%, n=2). This indicated that the selection of cases was an important aspect before the surgery.

Selection as the cause of poor outcomes was high because of the availability of vitro-retinal services. Cases with retinal diseases had cataract surgery mainly for better view during retinal surgeries. This might have been the reason for the higher selection of the cases. The poor outcome was related to surgical complications which occurred either intraoperatively or postoperatively even through the hands of experienced surgeons.

Postoperative sequelae mainly posterior capsular opacification was the cause in 2 of the patients resulting in poor vision. However, this would be treated subsequently with Nd: YAG (neodymium:yttrium-aluminum-garnet) capsulotomy. The poor visual outcome was noted due to surgical complications including the postoperative sequelae amounting to around 62.7% in the second RAAB survey (6). The vision of these patients did not improve with refraction because these patients already had pre-existing ocular pathology which hampered the final visual outcome. Therefore, the BCVA remained poor for these patients. The prospective assessment of surgical outcomes probably contributed to better surgical outcomes. Monitoring has enhanced the results of postoperative cataract surgery. The technique of tracking results and providing operating surgeons with feedback guarantees that the standard of postoperative eyesight is at the forefront and is constantly highlighted. The institution’s goal of recovering vision is always maintained, even if the operating doctors change, as is typical in any significant institution. Identifying the cases with any ocular pathology before the surgery would help plan and anticipate before surgery and also guide through patient’s counseling both preoperatively and postoperatively.

The strength of the study was that it was a hospital-based study (only a tertiary eye care center) and data collection was swift and suitable. The study included a large sample size which could likely be representative of the volume of SICS performed at the hospital and the study used a standard pro forma recommended by WHO.

A few limitations of the study were that it involved all the ophthalmologists at the Department of Ophthalmology who performed SICS and the conclusions from the study were drawn as a whole and not for individual surgeons. The study assessed outcomes only at 6 weeks post-SICS, providing insights into short-term outcomes. Longer-term follow-up data would have been valuable to assess the durability of visual improvements, refractive outcomes, and potential long-term complications. Although a considerable sample size was chosen, it was not representative when compared with other population-based studies. There was a loss in patient follow-up during the study period. The study was conducted at a single tertiary referral hospital, which might have limited the generalizability of the findings to other healthcare settings or populations.

## Conclusions

As a result of cataract surgery, patients with poor vision had improved visual acuity, according to this study. The results also showed how crucial a thorough pre-surgical assessment is to avoiding complications and guaranteeing a successful procedure. A good visual outcome was achieved, better than the WHO recommendation for tracking visual outcomes and in line with studies conducted in the area and other industrialized nations.

The success of cataract surgery should, however, be routinely monitored at hospitals in underdeveloped nations like Bhutan so that any challenges can be found and the proper steps can be made to close the gaps. To fully assess the total effect of cataract surgery on patients’ welfare and everyday functioning, this study advises long-term follow-up following cataract surgery.

## Data Availability

All Data are stored in the hard drive and can be made available on requirement and can be shared.

## Acknowledgment

I would like to express my gratitude to Dr. Nor Tshering Lepcha, Dr. Mendu Dukpa, and Dr. Karma Tenzin for their continued guidance throughout the planning, conduct, analysis, and manuscript writing of this thesis project. I would also like to thank Mr. Tshering Choeda and Mr. Indra Prasad Sharma.

I am thankful to the Faculty of Postgraduate Medicine, Khesar Gyalpo University of Medical Sciences of Bhutan (KGUMSB) and Jigme Dorji Wangchuck National Referral Hospital (JDWNRH) administration as well as to my fellow ophthalmology residents, optometrists, and ophthalmic assistants at Department of Ophthalmology.

I will remain forever indebted to all the consultants, patients, and their families who consented to participate in this study. I am grateful to the Ministry of Health for providing financial assistance to enable the smooth conduct of this study.

## References

1. Bourne RRA, Flaxman SR, Braithwaite T, Cicinelli M V., Das A, Jonas JB, et al. Magnitude, temporal trends, and projections of the global prevalence of blindness and distance and near vision impairment: a systematic review and meta-analysis. Lancet Glob Heal. 2017;5(9):e888–97.

2. Flaxman SR, Bourne RRA, Resnikoff S, Ackland P, Braithwaite T, Cicinelli M V., et al. Global causes of blindness and distance vision impairment 1990–2020: a systematic review and meta-analysis. Lancet Glob Heal. 2017;5(12):e1221–34.

3. National Statistical Bureau of Bhutan. Population Projections of Bhutan 2005–2030. http://www.nsb.gov.bt/publication/publications.php.id=2 Assessed on 07.06.2021.

4. Course S. Lens and Cataract. All about Your Eyes, Second Ed Revis Updat. 2021;74–81. Lepcha NT, Chettri CK, Getshen K, Rai BB, Bindiganavale Ramaswamy S, Saibaba S, et al. Rapid assessment of avoidable blindness in Bhutan. Ophthalmic Epidemiol. 2013 Aug;20(4):212–9.

5. WHO Informal Consultation on Analysis of Blindness Prevention Outcomes (1998: Geneva, Switzerland) & WHO Programme for the Prevention of Blindness and Deafness. (1998). Informal Consultation on Analysis of Blindness Prevention Outcomes, Geneva, 16–18. 1998.

6. Lepcha NT, Sharma IP, Sapkota YD, Das T, Phuntsho T, Tenzin N, et al. Changing trends of blindness, visual impairment and cataract surgery in Bhutan: 2009–2018. PLoS One. 2019 May 1;14(5).

7. Ferdosh JU, Uddin M, Husain R. Visual outcome after cataract surgery in a tertiary eye hospital of Chittagong district, Bangladesh. Asian J Med Biol Res. 2019;5(3):212–8.

8. Khanna RC, Rathi VM, Guizie E, Singh G, Nishant K, Sandhu S, et al. Factors associated with visual outcomes after cataract surgery: A cross-sectional or retrospective study in Liberia. PLoS One. 2020;15(5).

9. Cox JT, Subburaman GBB, Munoz B, Friedman DS, Ravindran RD. Visual Acuity Outcomes after Cataract Surgery: High-Volume versus Low-Volume Surgeons. Ophthalmology. 2019:126(11):1480–1489.

10. Matta S, Park J, Shantha GPS, Khanna RC, Rao GN. Cataract surgery visual outcomes and associated risk factors in secondary level eye care centers of L V Prasad Eye Institute, India. PLoS One. 2016;11(1):1–11.

11. Norregaard JC, Hindsberger C, Alonso J, Bellan L, Bernth-Petersen P, Black C et al. Visual outcomes of cataract surgery in the United States, Canada, Denmark, and Spain. Arch Ophthalmol. 1998;116(8):1095–100

12. Gogate P, Optom JJ, Deshpande S, Naidoo K. Meta-analysis to Compare the Safety and Efficacy of Manual Small Incision Cataract Surgery and Phacoemulsification. Middle East Afr J Ophthalmol. 2015;22(3):362–369.

13. Venkatesh R, Muralikrishnan R, Balent LC, Prakash SK, Prajna NV. Outcomes of high volume cataract surgeries in a developing country. Br J Ophthalmol. 2005;89(9):1079–83.

14. Manhas A, Manhas RS, Manhas GS, Gupta D. Visual Outcome among Patients Undergone Manual Small Incision Cataract Surgery Following their Identification in Screening Eye Camp in Jammu Province. Int J Contemp Med Res [IJCMR]. 2019;6(1).

15. Keel S, Xie J, Foreman J, Taylor HR, Dirani M. Population-based assessment of visual acuity outcomes following cataract surgery in Australia: The National Eye Health Survey. Br J Ophthalmol. 2018;102(10):1419–24.

16. Olawoye OO, Ashaye AO, Bekibele CO, Ajayi BG. Visual outcome after cataract surgery at the university college hospital, Ibadan. Ann Ib Postgrad Med. 2011;9(1):8–13

17. Bekibele CO, Fasina O. Visual outcome of traumatic cataract surgery in Ibadan, Nigeria. Niger J Clin Pract. 2008;11(4):372–5.

18. Hennig A, Kumar J, Yorston D, Foster A. Sutureless cataract surgery with nucleus extraction: outcome of a prospective study in Nepal. Br J Ophthalmol. 2003;87(3):266–70.

19. Kandel RP, Sapkota YD, Sherchan A, Sharma MK, Aghajanian J, Bassett KL. Cataract surgical outcome and predictors of outcome in Lumbini Zone and Chitwan District of Nepal. Ophthalmic epidemiology. 2010 Oct;17(5):276–81.

20. Bhatta S, Patel PJ, Awasthi S, Pant N, Pant SR. Visual outcomes of high-volume compared with low-volume manual small-incision cataract surgery in Nepal. Journal of Cataract & Refractive Surgery. 2020 Aug;46(8):1119–25.

21. Ruit S, Tabin GC, Nissman SA, Paudyal G, Gurung R. Low-cost high-volume extracapsular cataract extraction with posterior chamber intraocular lens implantation in Nepal. Ophthalmology. 1999 Oct;106(10):1887–92.

22. Yorston D, Gichuhi S, Wood M, Foster A. Does prospective monitoring improve cataract surgery outcomes in Africa? Br J Ophthalmol. 2002;86(5):543–7.

23. Hussen MS, Gebreselassie KL, Seid MA, Belete GT. Visual outcome of cataract surgery at Gondar university hospital tertiary eye care and training center, North West Ethiopia. Clin Optom. 2017;9:19–23.

24. Livingston PM, Lee SE, McCarty CA, Taylor HR. A comparison of participants with non-participants in a population-based epidemiologic study: the Melbourne Visual Impairment Project. Ophthalmic epidemiology. 1997 Jan;4(2):73–81.

25. Zafar S, Chen X, Sikder S, Srikumaran D, Woreta FA. Outcomes of resident-performed small incision cataract surgery in a university-based practice in the USA. Clinical Ophthalmology (Auckland, NZ). 2019;13:529.

26. Hashemi H, Mohammadi SF, Z-Mehrjardi H, Majdi M, Ashrafi E, Mehravaran S, Mazouri A, Roohipoor R, KhabazKhoob M. The role of demographic characteristics in the outcomes of cataract surgery and gender roles in the uptake of postoperative eye care: a hospital-based study. Ophthalmic epidemiology. 2012 Aug;19(4):242–8.

27. Murthy GV, Vashist P, John N, Pokharel G, Ellwein LB. Prevalence and vision-related outcomes of cataract surgery in Gujarat, India. Ophthalmic epidemiology. 2009 Dec;16(6):400–9.

28. Lam DS, Congdon NG, Rao SK, Fan H, Liu Y, Zhang L, Lin X, Choi K, Zheng Z, Huang W, Zhou Z. Visual outcomes and astigmatism after sutureless, manual cataract extraction in rural China: study of cataract outcomes and up-take of services (SCOUTS) in the caring is hip project, report 1. Archives of Ophthalmology. 2007 Nov;125(11):1539–44.

29. Gogate P, Vakil V, Khandekar R, Deshpande M, Limburg H. Monitoring and modernization to improve visual outcomes of cataract surgery in a community eyecare center in western India. Journal of Cataract & Refractive Surgery. 2011 Feb;37(2):328–34.

30. Khandekar RB, Jain BK, Sudhan AK, Pandey KP. Visual acuity at 6 weeks after small incision cataract surgery and role of audit in predicting visual acuity. European journal of ophthalmology. 2010 Mar;20(2):345–52.

31. Zhou B, Lu Y, Hajifathalian K, Bentham J, Di Cesare M, Danaei G, et al. Worldwide trends in diabetes since 1980: A pooled analysis of 751 population-based studies with 4.4 million participants. Lancet. 2016;387(10027):1513–30.

32. Yuan J, Wang X, Yang LQ, Xing YQ, Yang YN. Assessment of visual outcomes of cataract surgery in Tujia nationality in Xianfeng County, China. International Journal of Ophthalmology. 2015;8(2):292.

33. Kawasaki R, Wang JJ, Ji GJ, Taylor B, Oizumi T, Daimon M, Kato T, Kawata S, Kayama T, Tano Y, Mitchell P. Prevalence and risk factors for age-related macular degeneration in an adult Japanese population: the Funagata study. Ophthalmology. 2008 Aug;115(8):1376–81.

34. Varma R, Kim JS, Burkemper BS, Wen G, Torres M, Hsu C, Choudhury F, Azen SP, McKean-Cowdin R, Chinese American Eye Study Group. Prevalence and causes of visual impairment and blindness in Chinese American adults: the Chinese American eye study. JAMA ophthalmology. 2016 Jul;134(7):785–93.

35. Duan XR, Liang YB, Wang NL, Wong TY, Sun LP, Yang XH, Tao QS, Yuan RZ, Friedman DS. Prevalence and associations of cataract in a rural Chinese adult population: the Handan Eye Study. Graefe’s Archive for Clinical and Experimental Ophthalmology. 2013 Jan;251:203–12.

36. Henderson BA, Kim JY, Ament CS, Ferrufino-Ponce ZK, Grabowska A, Cremers SL. Clinical pseudophakic cystoid macular edema: risk factors for development and duration after treatment. Journal of Cataract & Refractive Surgery. 2007 Sep;33(9):1550–8.

37. Addagarla SR. Postoperative visual outcomes after cataract surgery in a teaching hospital. Indian Journal of Clinical and Experimental Ophthalmology. 2020;6(1):29–34.

38. Lundström M, Barry P, Henry Y, Rosen P, Stenevi U. Visual outcome of cataract surgery; study from the European Registry of Quality Outcomes for Cataract and Refractive Surgery. Journal of Cataract & Refractive Surgery. 2013 May;39(5):673–9.

39. Yorston D, Foster A. Audit of extracapsular cataract extraction and posterior chamber lens implantation as a routine treatment for age related cataract in east Africa. British Journal of Ophthalmology. 1999 Aug;83(8):897–901.

40. Egbert PR, Buchanan M. Results of extracapsular cataract surgery and intraocular lens implantation in Ghana. Archives of ophthalmology. 1991 Dec;109(12):1764–8.

41. Yorston D, Gichuhi S, Wood M, Foster A. Does prospective monitoring improve cataract surgery outcomes in Africa?. British Journal of Ophthalmology. 2002 May;86(5):543–7.

42. Signes-Soler I, Javaloy J, Montés-Micó R, Muñoz G, Albarrán-Diego C. Efficacy and safety of mass cataract surgery campaign in a developing country. Optometry and Vision Science. 2013 Feb;90(2):185–90.

43. Xiao B, Guan C, He Y, Le Mesurier R, Müller A, Limburg H, Iezze B. Cataract surgical outcomes from a large-scale micro-surgical campaign in China. Ophthalmic epidemiology. 2013 Oct;20(5):288–93.

44. Ezegwui IR, Ajewole J. Monitoring cataract surgical outcome in a Nigerian mission hospital. International ophthalmology. 2009 Feb;29:7–9.

45. Lai FH, Lok JY, Chow PP, Young AL. Clinical outcomes of cataract surgery in very elderly adults. Journal of the American Geriatrics Society. 2014 Jan;62(1):165–70.

46. Lepcha NT, Chettri CK, Getshen K, Rai BB, Bindiganavale Ramaswamy S, Saibaba S, Nirmalan PK, Demarchis EH, Tabin G, Morley M, Morley K. Rapid assessment of avoidable blindness in Bhutan. Ophthalmic epidemiology. 2013 Aug;20(4):212–9.

